# Risk factors for typhoid fever: A desk review

**DOI:** 10.1101/2024.09.10.24313440

**Authors:** Portia Boakye Okyere, Sampson Twumasi-Ankrah, Sam Newton, Samuel Nkansah Darko, Michael Owusu Ansah, Eric Darko, Francis Opoku Agyapong, Hyon Jin Jeon, Yaw Adu-Sarkodie, Florian Marks, Ellis Owusu-Dabo

## Abstract

**Background and Aim:** Typhoid fever, a significant global health problem, demonstrates a multifaceted transmission pattern. Knowledge of the factors driving the transmission of infection is critical for developing effective control strategies and resource allocation. This comprehensive desk review aimed at synthesizing evidence from 1928 to 2023 on risk factors associated with typhoid fever transmission.

**Method:** We conducted article searches in PubMed, Scopus, Google Scholar, and Semantic Scholar, using keywords related to risk, contributors, determinants, causes etc. associated with typhoid fever. We followed a registered protocol to support our search and triangulated the results.

**Results:** In all, we retrieved 1614 articles, of which 216 were reviewed. Of these articles reviewed, 106 provided data on typhoid fever risk factors. Unsurprisingly, of the total articles reviewed on risk factors, about 72% (76/106) originated from the Asian (48.1%, 51/106) and African (23.6%, 25/106) continents. A higher proportion, 47.2% (50/106) of the articles indicated risk factors related to socio-economic and housing transmission. Additional risk factors included foodborne transmissions (45.3%, 48/106), WASH: **Wa**terborne transmissions (42.5%, 45/106), **S**anitation and **H**ygiene practices (32.1%, 34/106), travel-related risk (16.0%, 17/106), antimicrobial agents (13.2%, 14/106), climate (13.2%, 14/106), environmental (8.5%, 9/106), typhoid carriers (10.4%, 11/106), and host risk (5.7%, 6/106) factors to disease transmission.

**Conclusion:** These findings highlight the necessity for targeted and combined interventions including improved sanitation infrastructure, enhanced WASH practices and the use of vaccines in endemic areas. Implementing effective strategies informed by this review can aid clinicians, public health experts, and policymakers in efficiently mitigating the burden of typhoid fever.

## Introduction

Typhoid fever (TF) is a potentially fatal febrile systemic disease caused by *Salmonella enterica* serotype Typhi (*S.* Typhi), a rod-shaped Gram-negative bacterium belonging to the *Enterobacteriaceae* family. *S.* Typhi exists exclusively in humans and causes illnesses that resemble many other febrile diseases^1^. A description of the infection was reviewed by Cunha^2^, clearly separating it from other febrile illnesses and associating its clinical manifestation with significant pathologic abnormalities in the spleen, mesenteric lymph nodes, and intestines. Nonetheless, the mainstay of diagnosis is a microbial culture, usually with blood or bone marrow samples. Although bone marrow culture is highly sensitive, it is both invasive and technically unfeasible in most settings. As a result, the disease is usually diagnosed with blood culture despite its limited sensitivity of about 40–80%, partly due to antibiotic exposures before the patient visits the health facility^3^.

The disease transmission is by the faecal-oral route and can take two main forms: direct transmission, where food and water in the immediate environment get contaminated through poor hygiene and sanitation practices, either by transient or chronic carriers; and indirect transmission, where the broader environment becomes contaminated when sewage pollutes water supplies, raw human faeces or untreated sewage are used as a crop fertilizer or piped water inadequately treated^4^.

Before 2000, the global burden of typhoid fever was estimated at 16 million illnesses and 600 thousand deaths annually^5^. In 2000, about 21.7 million illnesses and 216,000 deaths occurred^6^. By 2010, annual estimates indicated around 26.9 million cases and 200,000 fatalities^7^. A current estimate in 2017 suggests that the number of typhoid cases per year has decreased to 14.3 million, representing a 44.6% decrease in the global burden of typhoid fever^8^. Typhoid fever affects people of all ages, although children are more susceptible than other age groups^9^.

Typhoid fever can be prevented and controlled concurrently with vaccinations and advancements in food safety, water quality, hygiene and sanitation^10^. Three main generations of typhoid vaccines are presently approved for use by the World Health Organization (WHO): typhoid conjugate vaccine (TCV), live attenuated Ty21a, and the unconjugated Vi polysaccharide (ViPS) vaccines^4^. The WHO strongly recommends the use of TCV, for all ages due to its superior immunological properties, suitability for use in younger children, and predicted longer period of protection above 2 years, which was a major limitation for the use of the ViPS. However, to inform the choice of vaccination in a country, evidence on the scope of the problem as well as risk factors for the disease transmission in the area is required^10^. Despite notable progress in typhoid control, the disease remains a significant cause of morbidity and mortality to which billions of people worldwide are continuously exposed, particularly in Asia and sub-Saharan Africa.

The susceptibility of typhoid fever disease depends on a myriad of factors with distinct modes of transmission. In endemic countries, knowledge of typhoid fever risk factors is critical for developing effective control strategies and allocating resources. Several epidemiologic and modelled studies^11–13^ have explored location- and time-specific risk factors for typhoid fever. In addition, various review studies have been undertaken to comprehensively understand and address the risk of typhoid across different transmission routes. For instance, Lee et al.^14^ employed geospatial modelling to develop a typhoid risk index based on factors like water sources, toilet facilities, and population density, aiming to explain the geographical distribution of typhoid risk in impoverished countries. Likewise, Kim et al.^15^ modelled the incidence of typhoid by investigating the relationship between observed incidence rates and various geospatial covariates, such as access to improved water and sanitation, the health and environmental conditions of the population, which may influence the transmission of S. Typhi. Brockett and colleagues^16^ also systematically reviewed case-control studies to uncover associations between Water, Sanitation, and Hygiene (WASH) practices, food exposures, and typhoid fever. Similarly, Mogasale et al.^17^ performed a meta-analysis spanning 1990 to 2013 to estimate the risk of typhoid associated with inadequate access to safe water. Meanwhile, Ma et al.^18^ reviewed human genetic variants affecting susceptibility to enteric fever infection, and Levantesi et al.^19^ assessed the contribution of natural freshwater and drinking water as routes of *Salmonella* contamination from 2000 to 2010. While these studies collectively provide valuable insights into various aspects of typhoid risk, there remains a lack of a comprehensive review that synthesizes the existing knowledge on typhoid risk factors. This gap led us to ask: what do we know about the risk factors for typhoid fever? To address this, we aimed to conduct a desk review on risk factors for typhoid fever integrating the various transmission dynamics to inform targeted interventions for reducing typhoid incidence worldwide.

## Methods

### Search strategy

We searched PubMed, Scopus, Google Scholar, and Semantic Scholar for published articles on risk factors for typhoid fever. The search took place in June 2023 with titles and abstracts from databases downloaded and saved. Each database was searched using the following terms and keywords: Risk factors, factors, contributors, determinants, causes, predictors, susceptibility factors, factors of exposures, predisposing factor, typhoid fever, typhoid, *Salmonella* typhoid, *Salmonella* Typhi, *S.* Typhi, typhoid disease, typhoidal *Salmonellosis*, and typhoidal *Salmonella* and without perforation, complication, virulence, severe, and non-typhoid. We placed no restrictions on the publication year but language was restricted to English. We followed a protocol adapted from the PRISMA guidelines, which was registered with the Open Science Framework (OSF) on January 2024 (DOI: https://doi.org/10.17605/OSF.IO/87JV9), to structure our search. This study used published articles and as such, permission from the institutional review board was not required.

### Study screening and selection criteria

We screened and selected typhoid fever studies reporting on risk factors. In such studies, typhoid fever infections or outbreaks were required to be caused by *S.* Typhi either by culturing bodily fluids or stool, Polymerase Chain Reaction (PCR), Widal or any other serological test. Furthermore, we selected epidemiological or modelled studies of any design. Additionally, we excluded studies on severe infections (complication and mortality), studies that classified typhoid fever based on clinical indicators (i.e., signs and symptoms) or with unclear diagnostic methods and studies with non-human participants including animals, water samples, and farm-produce. Finally, articles with full text not written in English or cannot be located were excluded. The details of our inclusion and exclusion criteria can be located in the screener instruction section in the <u>review protocol</u>.

The titles and abstracts downloaded from each database were imported into EndNote X8.2 (Bld 13302), merged into a single reference list and duplicates eliminated. The de-duplicated list was then uploaded to an online systematic review tool Rayyan (Qatar Computing Research Institute, Doha, Qatar) for titles and abstracts screening^20^. Thereafter, all included citations were exported into Microsoft Excel, Version 16.16.27 (201012) for full-text retrieval and screening. Each subsequent process, including title and abstract review, full-text review, and data extraction, was performed by one author (Portia Boakye Okyere: PBO) with supervision from co-authors (Ellis Owusu-Dabo: EOD, Sam Newton: SN and Sampson Twumasi-Ankrah: STA). PBO resolved discrepancies collectively with the co-authors. Data were then extracted into Microsoft Excel and uploaded to a shared Google Sheets spreadsheet (Google LLC, Mountain View, CA, USA) (S2) and reported using Microsoft Word, Version 16.16.27 (201012). Co-authors reviewed the final dataset for completeness and accuracy.

### Data extraction

Electronic searches were performed using the internet to locate all eligible articles, and all relevant data relating to the research question were manually extracted into Microsoft Excel after reading the full text. The extracted data included specific risk factors for typhoid fever in all eligible articles. In addition, data on the route of transmission, sources of infection, year of publication, data collection period, town/district, country and continent of the study, study setting (outbreak or endemic), diagnostics method, study type, number of *S.* Typhi cases, total participants enrolled, ages of participants, study design, and citations were extracted (S2). We grouped the ages of participants into three based on inclusion age and age ranges/groups: “children” were ≤15 years, “adults” as >15 years, and “mixed ages” were both adults and children participants. Information on typhoid fever susceptibility was grouped according to their transmission routes; water-and food-borne transmissions, host risk factors, vaccination, travel-related risk, health education, occupational risk, population growth and overcrowding, sanitation and sewage systems, climate and meteorological factors, antimicrobial resistance, socio-economic and housing factors were extracted.

## Results

Our search strategy initially identified 1614 articles. After removing 217 duplicates (Figure 1), 1397 titles and abstracts remained for screening. Of these, 1181 were excluded, with majority (687 articles) not reporting typhoid fever risk factors as shown in Fig1. After a full-text review of 216 articles, 110 were further excluded, primarily due to the absence of clear risk factors in 34 articles. Other reasons for exclusion include 25 duplicates of the same study published by different authors in different journals, 10 studies involving non-human subjects such as farm-produced and water samples, 17 articles for non-English full-text and full-text not found, 6 articles not specifically related to *S.* Typhi, and 9 articles with missing or inappropriate diagnosis based on recall TF episodes, unclear diagnosis and clinical indicators (signs and symptoms). Furthermore, to avoid content duplication, 9 review articles were excluded. Finally, a total of 106 published articles were included in this review, ranging from^11–13,21–123^ (S2).

**Figure 1:**
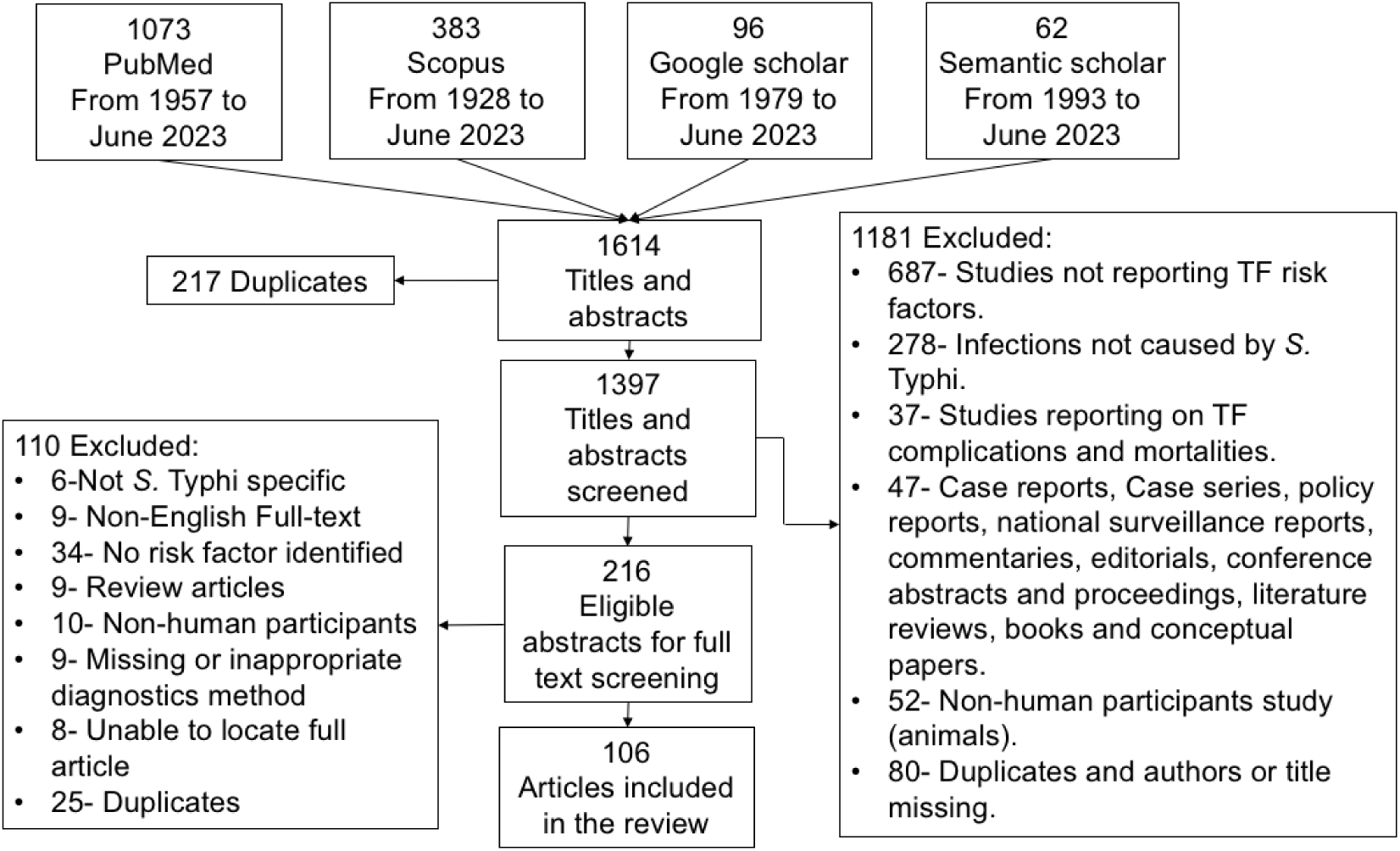
Desk review flow diagram of search strategies and selection of articles for risk factors for typhoid fever, in June 2023.

### Study characteristics

Among the 106 eligible articles (S2), data were collected from 1966 through 2020, spanning 6 continents: 23.6% (25/106) from Africa, 48.1% (51/106) from Asia, 0.9% (1/106) from South America, 8.4% (10/106) from North America, 9.4% (10/106) from Europe, and 6.6% (7/106) from Australia. Two articles (1.9%, 2/106) collected data from mixed continents, while one article (0.9%, 1/106) relied on the GeoSentinel Surveillance Network database, lacking specific location details (S1 Table 1). Regarding transmission routes, over 42.0% (45/106) of the articles examined typhoid fever risk factors suggesting waterborne transmission, while 45.3% (48/106) focused on foodborne transmission. Socio-economic and housing factors were covered in most (47.2%, 50/106) of the publications, hygiene and sanitation in 32.1% (34/106) articles, and all other TF risk factors in 74.6% (79/106) articles, (S1 Table 2). Across 101 papers, a total of 246,888 typhoid fever cases were reported, with a median (IQR) of 109 (50, 260). The diagnosis was confirmed predominantly through culture in 86.8% (92/106) of the articles, with 19.8% (21/106) articles using Widal and other serological tests and 5.7% (6/106) using PCR and other sequencing techniques. Age distribution data were available in 100 articles, with the majority 82% (82/100) including participants of mixed ages, while over 14% (14/100) focused on those ≤15 years and 4% (4/100) on those >15 years, (S1 Table 1).

## Discussion

Our desk review has allowed us to synthesize the evidence on typhoid fever risk factors from 1928 to 2023. We have identified specific risk factors suggesting waterborne and foodborne transmissions, socio-demographic, economic and housing factors, hygiene and sanitation, host risk factors, antimicrobial agents and typhoid carriers that could influence typhoid fever transmission. This is potentially important information for clinicians, public health experts and policymakers because effective control strategies can be developed as well as allocating limited resources mostly in endemic countries.

Approximately 72% of the reviewed articles collected data from Asia and Africa continents, where most of the developing countries are located. This is in accordance with the global burden of typhoid fever estimation, which shows that Asian and African countries bear the greatest burden^124^. The low number of articles in Europe, Northern America and Australia can be attributed to the introduction of control programs such as the treatment of municipal water, pasteurization of dairy products, and exclusion of human faeces from food production^1,125^. Likewise, the lack of studies from Southern America can be attributed to the decline in typhoid burden given the economic transition with improved water and sanitation in the area^1^.

### Waterborne transmission

Water sources greatly impact the spread of typhoid fever, with protected wells and piped water classified as safe, while rivers, streams and other unprotected sources are deemed unsafe^126^. Our review identified specific risk factors (Table 1) associated with typhoid fever and water sources, including having multiple drinking water sources^66^, use of non-municipal water sources for various purposes^27,66,105^, and main water sources with foul smell^35^. Regarding multiple water sources, households face potable water access challenges like limited hours, high tariffs, low pressure, and long distances from water sources. These challenges impede their ability to meet daily needs. Consequently, some households close to open-dug wells, rivers or streams may use these sources for bathing, cooking, or drinking^113^. Although convenient, these water sources often lack chlorination and may be contaminated with faecal matter, posing a risk for typhoid fever transmission. This risk is increased by foul-smelling water points that could contain pathogens such as *S.* Typhi^35^.

**Table 1.** Risk factors suggesting waterborne transmission of typhoid fever.

| Common factors | Specific risk factors |
| --- | --- |
| Water sources | Household sources of drinking water<sup>21,56</sup>, having multiple drinking water sources<sup>66</sup>, using water from other sources than the municipal water networks for bathing, brushing teeth or drinking<sup>110</sup>, primary water sources with unpleasant smell<sup>35</sup>, obtaining water from an outside tap<sup>44</sup>, unsafe water source<sup>51,113</sup>, obtaining water from municipal pipe for drinking<sup>105</sup>, obtaining water from a river/stream<sup>11,113</sup>. |
| Water supply | Water supplied by an outdated gravity-fed network<sup>51</sup>, Intermittent water availability<sup>11</sup>, inadequate safe water supply<sup>13</sup>, defective water systems<sup>111</sup>, household water supply from public wells and boreholes or merchants<sup>96</sup>, water from a community<sup>32</sup>, water from government overhead tanks<sup>41</sup>. |
| Contaminated/unsafe water | Drinking unsafe/contaminated water<sup>24,31,61,77,93</sup>, using substandard water<sup>81</sup>, *E. coli* in stored drinking water<sup>77</sup>, water sold in small plastic |
|  | bags<sup>78</sup>, use of ice cubes from a street vendor<sup>27,121</sup>. |
| Untreated water | Drinking water from a well<sup>94</sup>, untreated household drinking water<sup>35,44-46,68,118</sup>, use of untreated public water after rains<sup>30</sup>, drinking water from untreated open sources<sup>11,25,87</sup>, drinking water at the work site<sup>32</sup>, use of bore water<sup>105</sup>, Accidental ingestion of contaminated river water during swimming and/or bathing<sup>24</sup>, cooking and cleaning with river water or an open dug well<sup>66</sup>. |
| Water storage | Storing water in plastic containers without a lid<sup>96</sup>, Not storing water for drinking in a narrow-mouthed container<sup>36</sup>, not using tipped containers to draw water<sup>36</sup>, water sold in small plastic bags<sup>78</sup>. |

Defects in water supply systems can also facilitate the transmission of typhoid fever. Our study further discovered a typhoid fever outbreak associated with a gravity-fed network^51^, indicating a probable spread through an outdated mains system. Due to low water pressure, insufficient chlorination and faecal infiltration were implicated^11^.

Intermittent piped water supply, common in developing countries, poses health risks by causing contamination in non-pressurized pipes and creating negative pressure situations that enable pathogens like *S.* Typhi to enter the system, thereby increasing the risk of typhoid transmission^11,127^. Frequent interruptions in water supply also raise awareness of water storage practices, potentially increasing recontamination risks during storage and prompting households to turn to unsafe alternative water sources^11,27^. Households relying on water supplied from government overhead tanks^41^ and community or public taps^32,113^ may not always be microbiologically safe due to environmental conditions supporting contamination. Water contamination from various sources, including industrial activities, sewage discharges, agricultural practices, and urban runoff, poses other risks for typhoid transmission, as demonstrated by studies attributed to drinking contaminated water from unprotected groundwater sources and communal water sources exposed to human and animal waste^24,78^. The presences of *Escherichia coli* (*E. coli*) in drinking water is commonly regarded as an indicator of fecal contamination and a potential risk factor for typhoid fever due to the possible presence of enteric pathogens^77^. However, the relationship between *E. coli* and typhoid fever risk remains inconclusive. For instance, Karkey et al.^128^ observed a link between concentrations of *E. coli* in drinking water and the presence of *S.* Typhi nucleic acids. In contrast, Luby and colleagues^32^ found no significant difference in the levels of *E. coli* in household water samples between individuals with typhoid and control groups, challenging the consistency of this connection. Furthermore, water storage practices significantly impact typhoid transmission, with studies indicating a decline in microbiological water quality post-collection^96^. Increased contamination risk is associated with the type of vessel used to store or draw drinking water, with wide-mouthed containers posing higher infection risks compared to tipped containers due to their large open surface area and increased faecal contamination opportunities^36,96^. Untreated water, both inside and outside homes, poses significant risks for typhoid fever transmission^25,30,46,87^. Despite improvements in municipal water system filtration and chlorination, contamination with *S.* Typhi from wastewater cross-contamination remains possible^44^. Household water disinfection may be necessary to enhance water quality^35^. However, some households opt out of water treatment due to perceived safety^96^. Consuming water directly from pipes or other protected sources, even if contaminated, may contain insufficient bacteria to cause typhoid fever. Conversely, untreated water from unprotected sources may carry high *S.* Typhi levels, enough to cause clinical disease^24,32,45^. All risk factors discussed above are listed in Table 1.

### Foodborne transmission

Food serves as a highly efficient medium for the growth of *S.* Typhi compared to water^32^. This study revealed specific risk factors indicating potential foodborne transmission of typhoid fever (Table 2) with street stalls (restaurants) emerged as a major factor. Street food stalls are typically small, outdoor seating without refrigerators, easy access to potable water or adequate facilities for washing food and utensils^27,32,43^. Therefore, persons who frequently consume food from such establishments are at increased risk of developing typhoid fever^27^, possibly due to the use of untreated or tap water stored or served in contaminated containers for food preparation and drinking purposes^43^. Additionally, poor hygiene practices among street food vendors, including irregular handwashing^120^, and the potential exposure to carriers of *S.* Typhi contribute to the risk of infection^129^. Food preparers and handlers in the street or market eateries may lack knowledge of safe food production precautions to avert *S.* Typhi infections. They have no idea that they could be *S.* Typhi carriers because they did not apply for a license or registration from the food authorities, as a result, they are neither educated nor regularly inspected^43,106^. Consequently, they may share food and alcoholic beverages using poorly cleaned cups and other utensils between multiple clients^95,118^. Furthermore, Consumption of frozen foods such as ice cream^12,28,47^, shakes made from tropical fruits^82^ and iced drinks^27^ have also been linked to typhoid fever. Street vendors may purchase ice in large blocks made of unclean water from ice factories that produce ice for fisheries, not for human consumption, however, this ice is served in drinks for clients. Research has shown that *S.* Typhi can survive in ice for extended periods, thus the ice may potentially harbour the bacteria^27,43,120^. Likewise, the iced drink might be contaminated by street vendors who may be *S.* Typhi carriers in the distribution chain^27^. In contrast, dining at tea houses appears to decrease the risk of typhoid fever, as customers are primarily served boiled water and tea, limiting exposure to *S.* Typhi^45^. Consumption of uncooked or raw foods like onions, milk, meat, shellfish, papaya, cabbage and other traditional raw foods, poses a significant risk for typhoid fever^29,35,36,56,116^. Failure to wash fruits and vegetables before consumption also increases the risk of infection due to surface contamination^11,36,45^. Cross-contamination during meat handling may occur when the same knives and cutting boards for infected and uninfected meat are used in butcheries and restaurants. Furthermore, using contaminated water to wash carcasses and food contact surfaces might expose meat surfaces to *S.* Typhi^56^. Moreover, inadequate hygiene practices during milk processing may contribute to the contamination of dairy products, including butter, yoghurt, and homemade cheese^12,36,56^. While dairy animals do not harbour *S.* Typhi^130^, contaminated dairy products can serve as effective growth media for the pathogen^56^.

**Table 2.** Risk factors suggesting foodborne transmission of typhoid fever.

| Common factors | Specific risk factors |
| --- | --- |
| Street stall/restaurant food | Eating out from commercial food stalls/restaurants/mobile food vendors in the street/outside home<sup>23,32,43,89,99,120</sup>, eating outdoors at least once a week<sup>27,28,115</sup>, eating food from a roadside cabin during the summer<sup>32</sup>, mothers eating food from street vendors<sup>68</sup>, eating cold beverages outside home<sup>43</sup>, not dining at a tea-house<sup>45</sup>, consuming French fries with sauce and poppadum from street vendors<sup>105</sup>, eating commercially available foods or drinks<sup>118</sup>, eating food at community market<sup>95</sup>. |
| Poor food hygiene | Eating unwashed farm produced<sup>11</sup>, unwashed guavas<sup>36</sup>, not washing vegetables before eating<sup>45</sup>. |
| Food handlers | Consuming food items from vendors<sup>106,120</sup>, male food handlers<sup>38</sup>, consuming food with the mother's or caregiver's assistance<sup>28</sup>, drinking orange juice with hand contact<sup>48</sup>, poor/unhygienic food handling practices/procedures<sup>13,98</sup>, eating food prepared at home<sup>95</sup>. |
| Salads/other contaminated food | Eating contaminated foods<sup>61</sup>, including cucumber salad<sup>92</sup>, consumption of potato salad<sup>112</sup>, lettuce salad<sup>29</sup>, and raw salads (onion, cucumber, tomato)<sup>101</sup>. |
| Uncooked/raw food | Eating raw traditional foods like cig kofte<sup>29</sup>, raw onions and cabbage<sup>36</sup>, raw milk and meat<sup>56</sup>, uncooked shellfish<sup>116</sup>, intake of papaya<sup>35</sup>, unwashed guava<sup>36</sup>. |
| Frozen food | Eating of ice-cream<sup>28,32,47</sup>, ingesting ice cubes in beverages<sup>27</sup>, consumption of frozen tropical fruit (mamey) shakes<sup>82</sup>, fresh ice cream during the hot season<sup>12</sup>. |
| Milk products | Eating butter and yoghurt<sup>36</sup>, fresh yoghurt made from cow and sheep in the summer<sup>12</sup> and homemade cheese<sup>12</sup>. |
| Local/traditional food | Consuming locally-made beverages<sup>78</sup>, eating locally prepared popsicles<sup>101</sup>, consuming locally prepared flavoured beverages<sup>101</sup>. |
| Others | Eating food brought by relatives from endemic areas<sup>89</sup>, sharing food from the same plate<sup>121</sup>, sharing food plates<sup>51</sup>. |

### Socio-economic and housing factors

Our study findings presented in Table 3 showed socio-economic and housing factors as risks for typhoid fever transmission. Firstly, demographic characteristics including age and sex emerged as a major determinant of susceptibility to typhoid fever in this category. Gender dynamics introduce complexities, with conflicting findings regarding its association with typhoid fever risk. While a study suggests that males face greater exposure due to occupational, dietary factors or a lack of immunity^80^, others propose that females, influenced by their physiological status, hormonal imbalance and gender-specific activities, are more susceptible^118,119^. Nonetheless, a study by Rasul et al.^131^ maintains that typhoid fever incidence is independent of gender, affecting both males and females across all age groups equally. Age serves as a significant factor in typhoid transmission, with both young children and older adults identified as vulnerable groups^119,132^. Young children, characterized by their underdeveloped immune systems^31,40,85^ and limited understanding of disease transmission^56,85^, face heightened risks of infection. Whereas younger adults are predisposed to infections due to their adventurous lifestyle or unsanitary activities, such as eating junk food, and an increased number of social gatherings ^22,40,56^, older adults are more likely to be *S.* Typhi resistant due to continual immune-boosting^132^. Conversely, older adults may experience susceptibility as they are more exposed to occupational hazards of farming-related water contact activities^56,66^, rearing chicken or goats^66^, mishandling of *S.* Typhi samples by clinical staff^70,89^, and being a sewage worker^89^. With regards to rearing chicken, as *S.* Typhi is a human-adapted pathogen, it is likely that animal keeping is a confounder and may be associated with poor unhygienic conditions in the home^90^.

**Table 3.** Socio-economic and housing factors indicating typhoid fever risk.

| Common factors | Specific risk factors |
| --- | --- |
| Demographic background | Age<sup>22,56,85</sup>, Older age<sup>37,84</sup>, younger age<sup>31,40,57,80,106,107</sup>, sex of the individual<sup>100</sup>, being female<sup>106,119</sup> or male<sup>85,99</sup>, presence of preschool children in the household<sup>101</sup>, young adult<sup>119</sup>, demographic status<sup>54</sup> |
| Socio-economic status | Poor/low socio-economic status<sup>22,54,74,81,102</sup>, attending a gathering<sup>94</sup>, per capita disposable income of all residents, and per capita GDP<sup>100</sup>, unemployment or part-time work<sup>27</sup>, being part of a nuclear family<sup>47</sup>, wealth index<sup>88</sup>, being a student<sup>45</sup>, attending school or daycare<sup>66</sup> |
| Education level | No/low educational level<sup>25,46,118</sup>, educational level<sup>56</sup>, years of schooling<sup>88</sup>, students in conventional institutions of higher learning<sup>100</sup>, illiteracy rate<sup>47,102,103</sup> |
| Occupational risk | Mishandling of *S. Typhi* samples by clinical microbiology laboratory staff<sup>70</sup>, medical, laboratory personnel and sewage workers occupationally exposed to *Salmonella* bacteria<sup>89</sup>, household member growing crops<sup>66</sup>, farmers<sup>85</sup>, rearing chicken or goats<sup>90</sup>, job-related cause<sup>61</sup> |
| Population growth and overcrowding | Rising/bigger household size<sup>26,102</sup>, living in a crowded household<sup>29</sup>, crowding poor living conditions<sup>61</sup>, Increased population density<sup>31,111</sup> |
| housing system/ condition | Poor housing condition<sup>77</sup> |

Our investigation further revealed that socio-economic status correlates with an increased likelihood of *S.* Typhi infection^94^. Studies suggest that typhoid fever is more common in low-income countries and is connected to poor public health and low socio-economic indicators^22,81^. However, a contrasting study^88^ identified a protective effect of the wealth index on typhoid fever, suggesting that residing in affluent districts within low- or middle-income countries could significantly mitigate the risk. Moreover, the impact of per capita GDP and disposable income on typhoid fever transmission was identified^100^, highlighting the association between economic status and disease susceptibility. Hence, the likelihood of contracting typhoid fever may hinge upon various economic factors such as the economic foundation of the residency area, individual economic status, access to healthcare, and advancements in disease diagnosis, ultimately contributing to an increase in the detection rate of typhoid fever. Notably, insufficient case detection is observed when febrile patients fail to seek medical attention or undergo blood testing, emphasizing the critical role of consultation rates and diagnostic measures in controlling the spread of typhoid fever^100^.

In the context of education, our research highlights the lack of certificate education, indicating individuals who have never attended school, as a significant risk factor associated with the perception of typhoid fever. This suggests that a low level of education correlates with limited awareness of how the disease spreads^100,118^. Studies^88,103^ emphasize that the number of years spent in formal education directly influences one’s understanding of typhoid fever transmission. Specifically, uneducated people, exhibit a higher likelihood of contracting typhoid fever compared to those with formal education^102^. However, it is noteworthy that being a student^45^ and attending school (daycare)^66^ are independent risk factors for typhoid fever. Certain exposures within educational environments may increase the risk of infection among students. More importantly, health education, which encompasses awareness of practices such as handwashing with soap as recommended by WHO, is deemed essential. This form of education is considered a state of mind and may not necessarily be acquired solely through formal education systems. Table 5 illustrates specific risks associated with health education, such as a lack of knowledge regarding contact with typhoid fever^26,95^. Individuals lacking this knowledge are less likely to take preventive measures, increasing their susceptibility to infection.

Another significant factor for typhoid transmission (Table 5) aside from knowledge is the awareness of the presence of a typhoid patient at home ^118^. Individuals who are unsure of the presence of another/previous typhoid patient at home are more likely to have typhoid or a recurrence than those with full awareness. Typhoid patients may continue to shed *S.* Typhi in their stool and urine after initial antimicrobial treatment and up to 10% may do so for up to 3 months while others proceed to become asymptomatic carriers^1^. These transient or chronic carriers can be a source of reinfection^133,134^ or reactivation of previous infections^135^ at home and household members well informed of their presence may gain more knowledge about the disease, how it is transmitted, and how to avoid infection by the pathogen.

Recent contact with typhoid patients has also been observed as a risk factor in the Mekong Delta, southern Vietnam^46,86^. Traditional visits to sick persons might increase contact between individuals in the community. Close contacts who have visited a typhoid case are likely to live in the same area. Therefore, they might use the same water supply, i.e. transmission may still have occurred via water and not by person-to-person spread. Health education could assess and address the issue of contact with a sick person.

Lastly, overcrowding and housing conditions identified in Table 3 represent another critical dimension influencing typhoid fever transmission dynamics^29,31,102^. Overcrowding and poor housing infrastructure, particularly prevalent in squatter settlements, create environments conducive to person-to-person spread^26^. Inadequate access to sanitation facilities^13,111^ further exacerbates transmission risks, perpetuating cycles of infection within vulnerable populations.

### Hygiene and Sanitation risk factors

The risk factors outlined in Table 4 underscore the significant impact of hygiene and sanitation on the spread of typhoid fever. Poor handwashing practices emerge as a notable risk factor, given the crucial role hands play in transmitting *S.* Typhi through the oral-fecal cycle^23,28,118,121^. While handwashing with soap and clean water effectively removes pathogens^27^, inadequacies in technique, such as rinsing without soap^51,120^ or neglecting handwashing after defecation^11,95^, can increase the risk of bacterial spread^104^. Furthermore, using medicated soaps is an added advantage since it is more effective in eliminating bacteria from hands compared to regular soaps^26^.

**Table 4.** Hygiene and Sanitation risk factors for typhoid fever susceptibility.

| Common factors | Specific risk factors |
| --- | --- |
| Hygiene and Behavioral factors | Scarcity of soap near a hand washing facility<sup>26</sup>, non-use of soap for handwashing<sup>11,47,67,106</sup>, non-use of medicated soap<sup>26</sup>, non-availability of soap to wash hands after toilet use<sup>66,102</sup>, a habit of not washing hands before cooking/after defecating<sup>51</sup>, infrequent hand washing after latrine use<sup>11,95</sup>, poor hand washing practices<sup>103</sup>, occasionally/never washing hand with water and soap<sup>23,120</sup>, never/rarely washing hands before preparing/handling food, eating or feeding<sup>27,28,118</sup> |
| WASH practices | Poor WASH practices<sup>72</sup>, not living in a better WASH household<sup>73,104</sup> |
| Sanitation and sewage systems | Use of pit latrine<sup>25,90</sup>, open defecation<sup>90</sup>, improper disposal of solid waste<sup>25</sup>, burst sewer pipes at home<sup>94</sup>, living in houses with open sewers<sup>27</sup>, visible urine or faeces<sup>51</sup>, poor sanitary practice<sup>13,61,93,96,111</sup>, having home latrines<sup>36</sup>, no toilets at/in the residence<sup>106,113</sup>, poor excreta disposal<sup>77</sup>, having unimproved or malfunctioning sanitation infrastructure<sup>11</sup>, unsterilized water from the hospital disposal and residential sewage used to irrigate vegetable farmlands<sup>108</sup>, inadequate public sewerage system<sup>88</sup>, poor toilet drainage soil<sup>77</sup>. |

As mentioned earlier, typhoid fever is an oral route disease, and poor sanitation can trigger the disease^94^. The condition of the sewerage system has an important impact on typhoid fever incidence. Unimproved or damaged sanitation facilities and improper solid waste disposal are major sources of *S.* Typhi. According to Prasad et al.^11^, people lacking access to improved sanitation facilities or with damaged improved sewerage systems are particularly vulnerable. Moreover, household toilets built by persons without expertise are poorly constructed, situated in permeable soil, and prone to flooding and leaks, potentially leading to pollution of surface water and crops by human faeces^77^. Strengthening the installation of sanitary excreta facilities, along with proper waste disposal measures, is essential for more efficiently preventing typhoid fever. There is a direct correlation between improper solid waste disposal and the prevalence of typhoid fever^25^. Where sanitation and garbage disposal are lacking, typhoid fever continues to threaten life. Improper disposal of excreta within residential areas is associated with the risk of typhoid fever^77^. Preventing excreta from entering the domestic arena has a greater impact on typhoid transmission than behaviours preventing pathogens in the environment from being ingested (e.g. hand washing).

Furthermore, pit latrines, open defecation or sewer, burst sewer pipes at home and visible urine or faeces, have all been identified as risk factors for typhoid fever in several studies^27,51,90,94^.

Finally, the discharge of unsterilized water from hospital and residential sewage into rainwater canal systems used to irrigate farmlands where vegetables are cultivated leads to contamination of the produce^108^. These contaminated vegetables are often consumed without thorough washing, contributing to an increased incidence of typhoid fever. Particularly during the rainy season, when irrigation with more polluted water is common, the spread of pollution from farmlands, mixed with garbage, poses a heightened risk to human-inhabited areas.

### Other risk factors for typhoid fever

Table 5 presents other typhoid risk factors of antimicrobial agents, host factors, vaccination, travel-related risk, and environmental and climate conditions. Antimicrobial exposure has the greatest impact on *S.* Typhi infection. Prior use of antimicrobials at disease onset connects with an increased risk of typhoid infection, particularly concerning both multiple (MDR) and extensively (XDR) drug-resistant strains^61,79^. Likewise, studies indicate a relationship between frequent or recent antimicrobial use within four weeks preceding illness onset and the occurrence of typhoid fever ^32,42,45,99^. Antimicrobial exposure can induce prolonged alterations in gut flora and compromise the barrier against bacterial colonization, thereby reducing the threshold of *S.* Typhi required for infection ^45^. Furthermore, we discovered in studies such as Yousafzai et al.^117^, Srinivasan et al.^101^, and Kamal et al.^79^ emphasizing antimicrobial resistance as a risk factor for typhoid fever with certain resistant strains capable of causing epidemics. Antimicrobial resistance is caused by the routine presence of *S.* Typhi in the human intestine, and indiscriminate antibiotic usage^42,61^. Consequently, drug-resistant *S.* Typhi strains, facilitated by the presence of multiple virulence factors, are becoming more frequent worldwide. Notably, we identified specific risk factors associated with *S.* Typhi-resistant strains encoding a range of virulence genes, including those within the H58 lineage, capable of infecting and interacting with host cells^58,60,101,119^.

**Table 5:** Other risk factors for typhoid fever.

| Common factors | Specific risk factors |
| --- | --- |
| Antimicrobial Agent | Frequent/history/ of antimicrobial usage<sup>32,45,53,60,61,74,79,99</sup>, Chloramphenicol-resistant *S. Typhi* strain<sup>42,77</sup>, ceftriaxone-resistant *S. Typhi* strain<sup>117</sup>, MDR or XDR *S. Typhi* strains<sup>119</sup>, circulation of virulent *S. Typhi* strain (H58-lineage)<sup>58,101</sup>. |
| Host risk factors | Polymorphism in intronic VNTR of IL-4<sup>33</sup>, presence of serum anti-*H. pylori* IgG antibodies<sup>47,107</sup>, history of chronic underlying disease<sup>67</sup>, HIV infections<sup>69</sup>, Haplotype of TNF locus from SNPs<sup>122</sup> |
| Typhoid carrier/s | Recent/close contact with confirmed/active typhoid fever patient<sup>30,46,61,86,99,103</sup>, hospitalization of household member with febrile illness<sup>66</sup>, history of typhoid fever infections<sup>67</sup>, having typhoid carrier at home<sup>63</sup>, recent typhoid fever case in the household<sup>106</sup>, having a housekeeper (a boy or girl)<sup>118</sup>. |
| Vaccination | No/lack of vaccination<sup>61,73,104</sup>, vaccine hesitancy<sup>118</sup>, vaccine ineffectiveness<sup>92</sup>, poor vaccination coverage<sup>101</sup> |
| Health education | Lack of knowledge regarding typhoid fever contact<sup>26</sup>, poor awareness of typhoid fever disease<sup>95</sup>. |
| Travel-related risk | Longer duration of stay in the endemic area<sup>37</sup>, returning/visiting endemic countries<sup>74,84,114</sup>, visiting friends and relatives in endemic areas<sup>37,71</sup>, travel destination<sup>62</sup>, travel outside the United States or Sweden (international travel)<sup>39,57,91</sup>, Asian travellers<sup>50</sup>, children visiting friends and relatives in endemic places (particularly South Asia)<sup>59,97</sup>, recent travel to endemic areas<sup>83,123</sup>, transient male workers<sup>80</sup>, living in a metropolitan area<sup>84</sup>, urbanization<sup>88</sup>, number of foreign tourists received (tourism)<sup>100</sup> |
| Environmental conditions/factors | Living in geographically lower elevation areas<sup>40</sup>, neighbours to a typhoid fever case<sup>52</sup>, potentially floodable areas<sup>55</sup>, proximity to major rivers and creeks<sup>55</sup>, housing (external condition)<sup>77</sup>, a lack of agricultural land<sup>113</sup>, hydrological catchment areas<sup>119</sup>, residing closer to waterbodies, residing near typhoid study treatment centres<sup>102</sup>, Anthropogenic alteration of land cover and hydrology<sup>76</sup>, Environmental factors<sup>54</sup> |
| Climate/<br>Meteorological factors | Seasonal variation or fluctuations<sup>22,80,100,108</sup>, high temperatures during summer<sup>101</sup>, rainfall<sup>55,113</sup>, temperature and precipitation<sup>109,113</sup>, high vapour pressure<sup>113</sup>, rainy and harmattan seasons<sup>34</sup>, extreme weather conditions<sup>75</sup>, higher/hot temperatures<sup>64,84</sup>, flooding<sup>85</sup>, wind speed<sup>88</sup> |

### Host risk factors

Host genetic factors influence susceptibility to infectious diseases in humans. Our review turned up a study by Mustafa et al.^33^ that explored the relationship between genetic polymorphisms and the risk of typhoid fever. The findings suggested that individuals with the 2R3R heterozygote for the intronic VNTR of IL4 may be genetically predisposed to typhoid fever. However, a study by Dunstan et al.^122^ suggested that a haplotype in the TNF locus offers protection from typhoid. Another study^47^, found a link between serum anti-H. pylori immunoglobulin G (IgG) antibodies and an increased risk of typhoid fever. The presence of serum IgG antibodies indicates either prior or active infection with H. pylori, as antibodies can persist even after infection clearance, potentially increasing the risk of infections^47,136,137^. A possible explanation is that the gastric-acid barrier serves as a crucial defense mechanism against *S.* Typhi infection. H. pylori infection has been associated with hypochlorhydria, a condition that impairs the gastric-acid barrier in the stomach^47,107^. This impairment may be the mechanism through which H. pylori infection increases the risk of typhoid fever.

A substantial relationship between typhoid fever and having a chronic underlying condition disease was also identified in our review^67^. This could be explained by the fact that *S.* Typhi was not cleared from the body due to the weakening immune system caused by the underlying chronic diseases. For instance, while *S.* Typhi is not widely associated with AIDS in developed countries, this relationship is observed in endemic areas. We uncovered a study by Gotuzzo et al.^69^ that found an increased risk of typhoid in HIV-infected patients from typhoid-endemic areas. It is worth mentioning that the majority of the HIV-positive patients enrolled in the study were homosexuals, suggesting a potential increase in typhoid incidence due to direct faecal-oral transmission among the homosexual population.

### Travel-related risk

Typhoid fever, once prevalent in industrialized countries, is now effectively controlled^89,91^, yet imported infections remain a significant public health concern^37,50,83^. In our study, we identified the risk of typhoid fever associated with travel, as detailed in Table 5. The risk of infection among travellers varies depending on factors such as age, destination, duration, and purpose of travel^97^. Travellers visiting friends or relatives (VFRs) are in a high-risk category for typhoid fever^37,59,71^. Because they are much less likely than other travellers to seek pre-travel counselling, they may visit more rural, remote areas and engage closely with local people as well as eat high-risk foods and beverages^49^. Children and young adolescents who are VFRs are also at high risk of contracting typhoid fever due to a lack of immunity or the possibility of travelling under unhygienic conditions^59,80,97^. We also discovered that travelling to endemic locations increases the risk of contracting typhoid^50,57,84,91^. According to Lin et al.^84^, over half of all travellers with typhoid returning to developed countries come from Asia or Africa, where the disease is widespread. This may also reflect increased exposure to these environments. Moreover, the risk of infection is highest during extended journeys to endemic areas. Short-term visitors to such areas face lower risk^37^. Economic globalization facilitates the movement of travellers for business or tourism, contributing to disease spread. Consequently, the influx of workers and foreign tourists, often without proper disease-preventive education or vaccines, increases the risk of typhoid infection^80,100^.

### Vaccination

Vaccination is essential for the control of typhoid fever in endemic and epidemic settings as well as among travellers moving between non-endemic and endemic areas. The WHO recommends the programmatic use of typhoid fever vaccines in endemic areas^4^. We retrieved two studies^73,104^ where the risk of typhoid fever was reduced among populations who had received effective typhoid vaccines and lived in households with better water, sanitation, and hygiene (WASH) conditions, Table 5. Conversely, poor vaccination coverage or the absence of vaccination with suboptimal WASH can exacerbate typhoid transmission in a given area^61,101^. Defective vaccine efficacy can result from various factors, including faulty batches, incorrect immunization procedures, or the interval since vaccination. In our study, we found evidence of ineffective vaccines increasing the risk of typhoid fever, particularly in individuals vaccinated more than three years prior^92^. This is due to vaccine weaning as polysaccharide vaccines exhibit a cumulative efficacy of approximately 55% over three years, with significant protection observed primarily within the first two years post-immunization.

### Environmental, seasonal and climate factors

Typhoid fever transmission exhibits distinct seasonal patterns influenced by environmental and climatic factors such as temperature, humidity, and precipitation. In this study, diverse peak risk periods have been identified in different endemic regions. While Taiwan registers a surge in cases during the fall (September-November) and winter seasons (December-February)^84^, in India, Allahabad experiences its peak in June^22^, with Ahmedabad seeing a spike during the monsoon season (July-November)^75^. Another study by Corner et al.^54^ discovered that almost half of the yearly typhoid cases, reaching up to 11 per 100,000 individuals in the Dhaka Metropolitan Area, occurred during summer and fall (July-October). Additionally, Srinivasan and colleagues^101^ found a correlation between hot weather (summer i.e. June-August) and increased typhoid cases. These fluctuations stem from a complex interplay of climatic conditions, hygiene practices, and local cultural dynamics of an area^84,138^. For instance, during summer seasons or in regions with warmer temperatures, *S.* Typhi multiplication in contaminated foods is enhanced^101^, likewise, during colder temperature and humidity conditions, *S.* Typhi survive longer in water and soil^139^. These changes in climatic conditions together with increased human movement interact to drive typhoid fever infection in many endemic areas. Furthermore, heavy rainfall can trigger flooding, leading to the contamination of water sources through sewage overflow^22,64^ exacerbating typhoid incidences. Similarly, increased precipitation associated with the rainy season may facilitate the spread of *S.* Typhi through water and food contamination^75^. In areas where surface water is relied upon for daily needs, such as low-lying areas, the risk is compounded during the rainy season due to sewage runoff^40,54,55^. Settlements situated in flood-prone or hydrological terrains, like river floodplains, face severe typhoid transmission risks, particularly during the rainy season^65,76^.

## Conclusion

Identifying the risk factors contributing to typhoid fever is crucial for understanding and controlling its transmission. Effective prevention and control require addressing these factors by improving water and food quality, enhancing hygiene and sanitation, implementing mass vaccination programs, and promoting health education. Interventions targeting environmental and climate-related transmission are also essential for mitigating the risks associated with typhoid fever.

### Study limitation

Risk of bias was not assessed and this study is limited to English language articles.

## Data Availability

This study used published articles and all data collected from included articles are compiled and attached as a supporting information (S2_file).

## Acknowledgement

Special thanks to the EOD projects at Kwame Nkrumah University of Science in collaboration with the International Vaccine Institute for their support.

## Author contributions

The study was conceived by EOD and PBO, with PBO leading the development of the review protocol. EOD, SN, and STA reviewed the protocol before its registration with OSF. PBO conducted the literature search, screened titles and abstracts, reviewed full texts, and performed data extraction. Any discrepancies were resolved collaboratively by EOD, SN, and STA, who also reviewed the final dataset. PBO and STA carried out data analyses. All authors contributed to the manuscript’s drafting, reviewing and editing for publication. All authors agreed and approved the final manuscript for publication.

## Supporting information

**S1 Table 1.** Summary of study characteristics

**S1 Table 2.** Distribution of transmission routes and common risk factors for typhoid

**S2.** Data extraction entities to: Risk factors for typhoid fever, a desk review.

